# The Impact of Fasting the Holy Month of Ramadan on Colorectal Cancer Patients and Two Tumor Biomarkers: A Tertiary-Care Hospital Experience

**DOI:** 10.1101/2022.08.04.22278413

**Authors:** Kanaan Alshammari, Haifa Alhaidal, Reem Alharbi, Alanood Alrubaiaan, Ghadah Alyousif, Mohammad Alkaiyat, Wesam Abdel-Razaq

**Affiliations:** King Abdulaziz Medical City, Ministry of National Guard Health Affairs, Riyadh, Saudi Arabia; College of Pharmacy, King Saud bin Abdulaziz University for Health Sciences, Ministry of National Guard Health Affairs, Riyadh, Saudi Arabia; College of Medicine, King Saud bin Abdulaziz University for Health Sciences, Ministry of National Guard Health Affairs, Riyadh, Saudi Arabia; King Abdullah International Medical Research Centre, Ministry of National Guard Health Affairs, Riyadh, Saudi Arabia

**Keywords:** Intermittent Fasting, Colorectal Cancer, Tumor Biomarkers, Carcinoembryonic Antigen (CEA), Lactate Dehydrogenase (LDH)

## Abstract

**Background:** Fasting during the holy month of Ramadan is a religious ritual practiced by the majority of Muslims around the globe. This daytime fasting is short-term or intermittent fasting, which may be associated with valuable health benefits, particularly in cancer patients.

**Methods:** A prospective cohort study of pre-and post-evaluation of 37 colorectal cancer (CRC) patients was conducted in the oncology outpatient clinics to assess the impact of fasting during Ramadan on the tolerability of chemotherapy side effects and to assess changes in the levels of carcinoembryonic antigen (CEA) and lactate dehydrogenase (LDH) tumor biomarkers.

**Result:** The vast majority of CRC patients (89.2%) had fasted at least part of the month of Ramadan. Most (73%) reported “Serenity” after fasting during Ramadan with improved tolerability of chemotherapy side effects. The results did not reveal any significant difference in the measured laboratory variables between pre-fasting values and by the end of the 30 days of Ramadan. Although statistically insignificant, the levels of CEA and LDH were reduced in 46.9% and 55.6% of CRC patients, respectively. The mean level of CEA in the fasting group was substantially reduced by more than 40%, attributed to the highly significant decline of CEA levels in three patients only (p=0.0283).

**Conclusion:** The current study confirms the safety and tolerability of intermittent fasting in CRC patients actively receiving chemotherapy, which is consistent with several reports. Nonetheless, the results did not reveal a significant decrease in CEA and LDH tumor biomarkers.

## Introduction

Daytime fasting during the month of Ramadan is a religious ritual practiced by the majority of Muslims around the globe. Ramadan is the ninth month of the lunar, also known as the Hijri calendar. Ramadan fasting is considered a distinctive model of intermittent fasting that refrains eating and drinking all forms of foods, beverages, and even oral medications from sunrise to sunset each day until the end of the month of Ramadan, which may extend for 29 or 30 days depending on the visibility of the crescent moon. Such changes in the daily diet pattern may dramatically affect individual metabolic functions [1]. Several studies have reported valuable beneficial outcomes for intermittent fasting, including controlling obesity [2], energy metabolism [3], blood pressure [4], diabetes [5], cancer [6], and other diseases. Although Muslims worldwide are fasting the holy month of Ramadan on an annual basis, the health consequences of such lifestyle practices are not adequately studied. Therefore, it is pivotal to evaluate the safety of this fasting practice in individuals with compromised health conditions, particularly cancer patients who are receiving cytotoxic chemotherapy.

In the Gulf countries, including Saudi Arabia, colorectal cancer (CRC) is still the topmost commonly diagnosed cancer in men and the third most common cancer, after breast and thyroid cancer, in women [7]. Although CRC is usually diagnosed in the elderly population, recently, the incidence rate has peculiarly increased in younger adults under 50 years [8] [9]. Several therapeutic modalities were employed in treating CRC patients, including surgery, radiation, and systemic chemotherapy, primarily depending on the cancer stage and patient’s factors. Chemotherapy regimens were shown to be favorably effective in advanced CRC stages [10]. However, cytotoxic chemotherapies have always been associated with numerous toxicities and adverse events. These include, but are not limited to, nausea and vomiting, myelosuppression, cardiotoxicity, renal impairment, fatigue, peripheral neuropathy, and others.

This study assessed the impact of fasting during the holy month of Ramadan on CRC patients concerning their tolerability of chemotherapy side effects, besides changes in blood parameters and levels of two tumor biomarkers [namely, carcinoembryonic antigen (CEA) and lactate dehydrogenase (LDH)], which are primarily associated with certain types of carcinomas, including CRC.

## Methods

### Study design

A prospective cohort study of pre-and post-medical and clinical evaluation of histologically confirmed CRC patients was conducted in the oncology outpatient clinics at King Abdulaziz Medical City (KAMC) and King Abdullah Specialized Children Hospital (KASCH), Riyadh, Saudi Arabia. A total of thirty-seven (37) CRC patients were followed from the beginning of the month of Ramadan until the 20th of the next Hijri month of Shawal. Patients’ demographic information including age, gender, weight, height, health status, and the received chemotherapies were retrieved from patients’ electronic medical records. Baseline clinical data, including complete blood count, renal and liver functions, and serum level of two tumor biomarkers (CEA and LDH) were measured immediately before the beginning of the fasting month of Ramadan and by the end of the follow-up period. After obtaining their informed consent, all patients were asked to fill in a mini questionnaire-based survey that explored their practice and attitudes towards fasting during the month of Ramadan, whether they complied with healthcare providers’ recommendations, and their subjective responses regarding tolerance of chemotherapy adverse effects.

### Statistical analysis

Results are expressed as mean ± standard deviation (SD) and median with interquartile range (IQR) as dispersion characteristic for continuous data. Categorical variables are represented as proportions of total contributors. Graphs and statistical analyses were performed using GraphPad Prism® software package version 9.0 (San Diego, CA, USA). Statistical significance was considered at p-values less than 0.05 using Unpaired Student’s t-test between groups or Wilcoxon matched-pairs Student’s t-test” within the same group.

## Results

A total of 37 CRC patients’ data were reviewed. Table-1 displays the general characteristics of enrolled patients. The mean age was 52.7 ± 11.6 years, with a median value of 53 years (range 30-76). Almost half of the sample (45.9%) were overweight or obese patients with body mass indices of more than 25. Most of the sample had advanced stage III or IV carcinomas (27.0% and 67.6%, respectively). Most patients were treated with 5-fluorouracil-based chemotherapies (FOLFOX or FOLFIRI regimen, that included fluorouracil and leucovorin added to either oxaliplatin or irinotecan) as the standard of care treatment of advanced CRC. A bit more than 50% of patients also received a biological targeted therapy (Cetuximab n=8, Bevacizumab n=8, and Panitumumab n=2) in addition to their primary standard cancer chemotherapy.

**Table-1:** General profile and cancer characteristics of enrolled CRC patients, n = 37

| Variable | Value |
| --- | --- |
| Gender n (%) |  |
| Male | 17 (45.9%) |
| Female | 20 (54.1%) |
| Age (in years) |  |
| Mean ± (SD) | $52.7 \pm (11.6)$ |
| Median (range) | 53 (30-76) |
| BMI n (%) |  |
| < 18.5 (underweight) | 4 (10.8%) |
| 18.5-24.9 (healthy weight) | 16 (43.2%) |
| 25-29.9 (overweight) | 9 (24.3%) |
| ≥ 30 (obesity) | 8 (21.6%) |
| Smoking n (%) |  |
| No | 35 (94.6%) |
| Yes | 2 (5.4%) |
| Hypertension n (%) |  |
| No | 27 (73.0%) |
| Yes | 10 (27.0%) |
| Hyperlipidemia n (%) |  |
| No | 30 (81.1%) |
| Yes | 7 (18.9%) |
| Diabetes n (%) |  |
| No | 21 (56.8%) |
| Yes | 16 (43.2%) |
| CA Family History n (%) |  |
| No | 33 (89.2%) |
| Yes | 4 (10.8%) |
| ECOG Performance Status n (%) |  |
| 0 | 13 (35.1%) |
| 1 | 22 (59.5%) |
| 2 | 2 (5.4%) |
| 3-5 | 0 (0.0%) |
| Tumor site n (%) |  |
| Colon | 27 (73.0%) |
| Rectosigmoid | 1 (2.7%) |
| Rectal | 9 (24.3%) |
| Cancer Stage n (%) |  |
| II | 2 (5.4%) |
| III | 10 (27.0%) |
| IV | 25 (67.6%) |

Patients were asked to fill in a mini-questionnaire survey exploring their fasting practice during the month of Ramadan (data not shown). About 92% of patients had consulted with their healthcare providers about the safety and appropriateness of fasting during the month of Ramadan, which coincided with their chemotherapy cycles. Almost all healthcare providers assured the safety of such intermittent fasting on CRC patients. Nearly one-third of patients had asked for dose rescheduling after sunset so that it would not affect their fasting practice during the daytime. In general, most patients (73%) reported “Serenity” after fasting during the holy month of Ramadan with perceived improved tolerability of chemotherapy side effects. About 8% of patients had unplanned visits to the emergency room during the month of Ramadan due to either pain or fever; one patient reported a skin rash. Less than 20% of responders reported feeling nauseous while fasting, but it did not affect their activities of daily living in most patients. This was confirmed by the Eastern Cooperative Oncology Group (ECOG) performance status scale, which determines patients’ ability to carry out their daily living activities during cancer chemotherapy and is reported by the treating physician (for details, see Table-1).

The vast majority of patients (89.2%, n=33) had fasted at least part of the month of Ramadan. On average, patients had fasted for 25 ± (5.3) days (range 10-30 days). Thus, it was decided to categorize patients into; Group A, which included patients who did not fast (n=4, 10.8%) or had fasted less than 20 days (n=7, 18.9%), and Group B, those patients who had fasted the entire 30 days of the month of Ramadan (n=13, 35.1%) or had fasted at least 20 days (n=13, 35.1%). Table-2 presents various demographic and clinical data of patients in both groups. The two groups show no significant difference, implying similar initial variables.

**Table-2:**
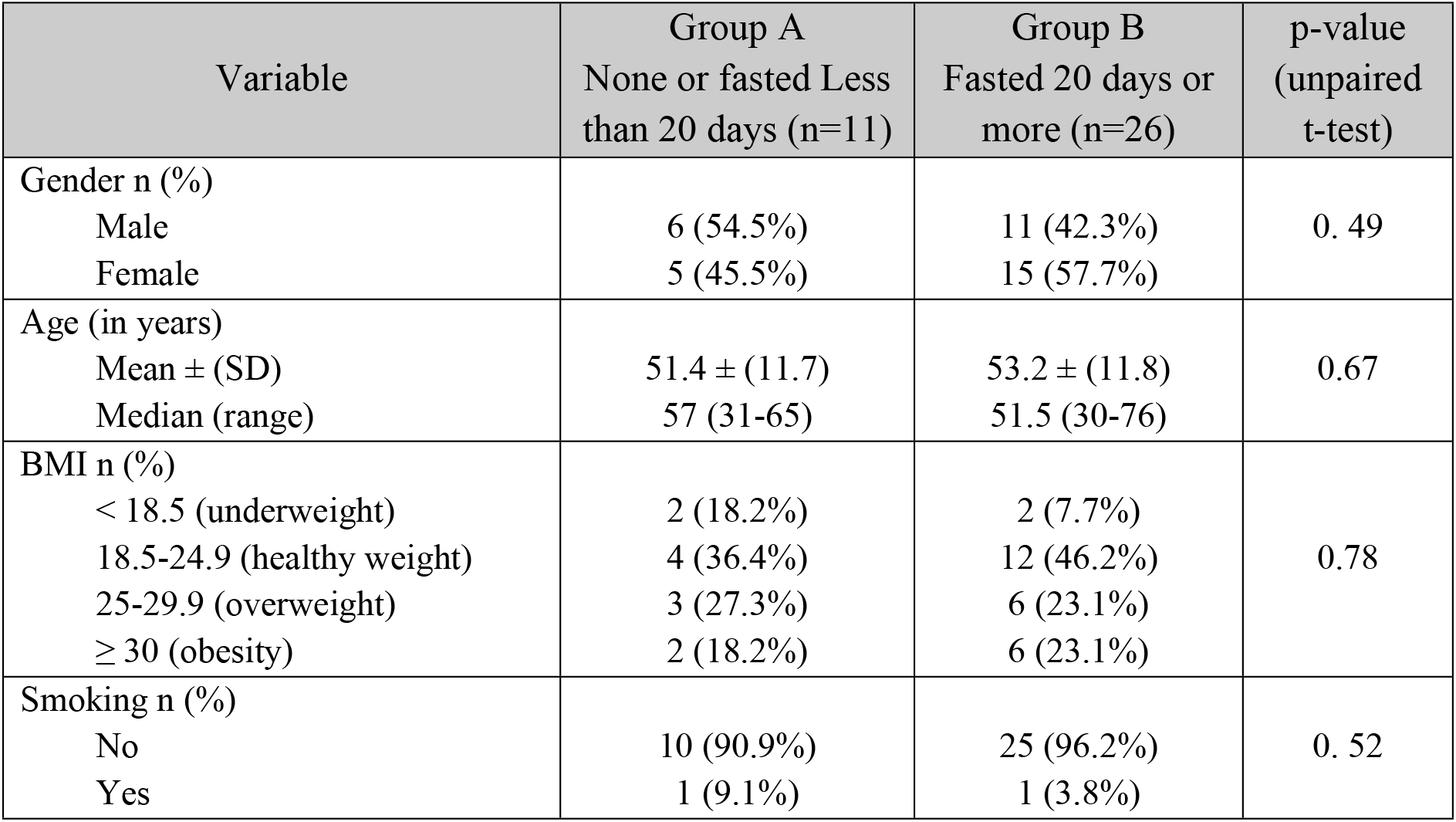
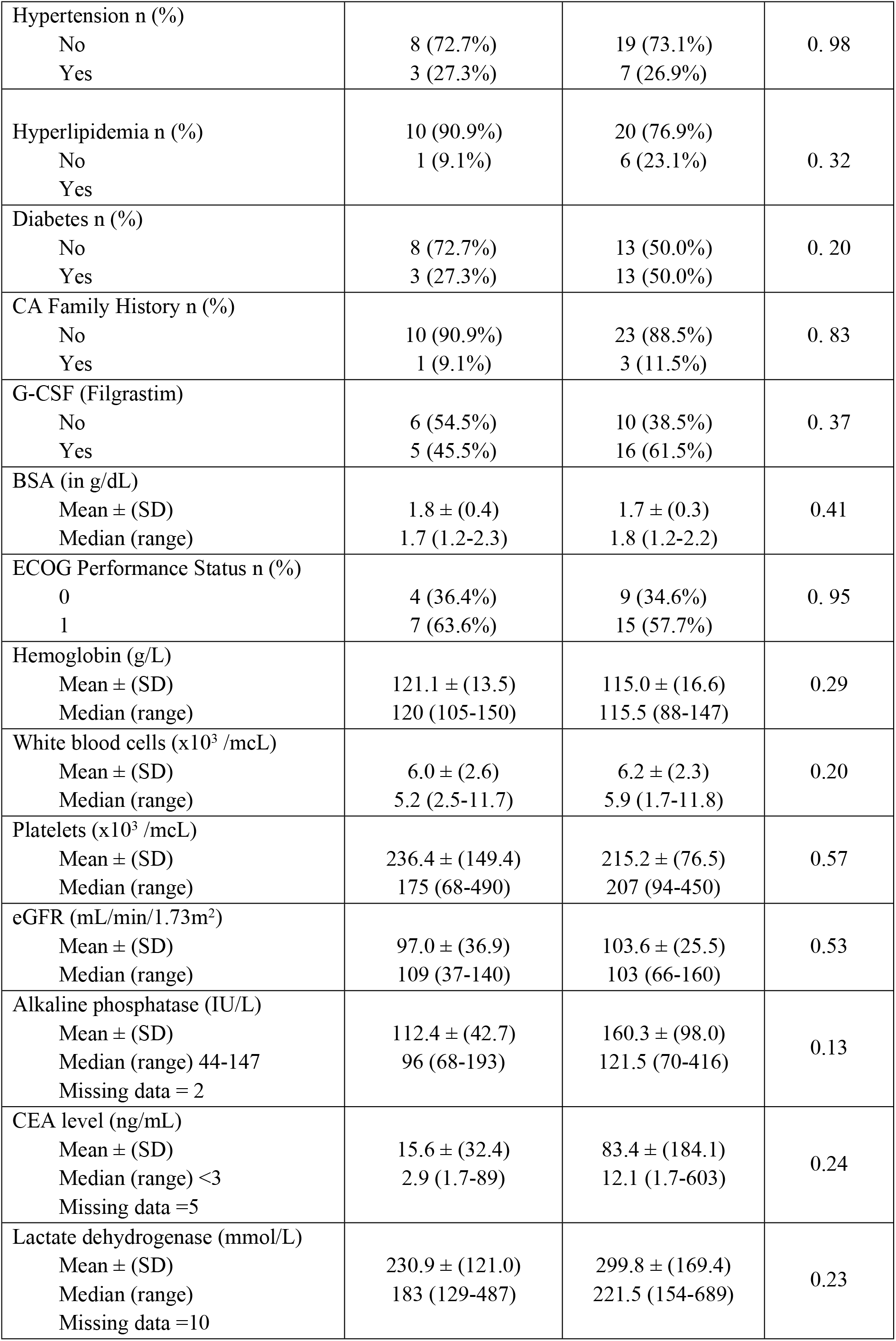
Initial variables among the low-fasting and the high-fasting groups, n = 37

On the other hand, Table-3 shows the mean differences between initial variables and after fasting, if any, by the end of the 30 days of the month of Ramadan. The measured blood parameters were almost comparable. Although statistically insignificant, both tumor biomarkers, CEA and LDH, were reduced by 12.4% and 21.8%, respectively, in Group-A; and declined by 40.9% and 15.5%, respectively, in Group B. Nevertheless, one sample in group A showed a noteworthy decrease in CEA levels from 89 to 54 mmol/L, whereas another sample in group-A showed an increase in CEA levels from 6.9 to 29.2 mmol/L (See Figure-1). Three samples in Group B showed momentous decreases in CEA levels from a mean value of 568.3 ± (32.7) to 296.0 ± (103.6) ng/mL with a highly significant p-value of 0.0283.

**Table-3:** Mean difference value analysis, n = 37

| Fasting | Group A:<br>None or fasted less than 20 days<br>(n=11) |  |  |  | Group B:<br>Fasted 20 days or more<br>(n=26) |  |  |  |
| --- | --- | --- | --- | --- | --- | --- | --- | --- |
|  | Before<br>Mean<br>± (SD) | After<br>Mean<br>± (SD) | Mean<br>Diff<br>(%) | p-value* | Before<br>Mean<br>± (SD) | After<br>Mean<br>± (SD) | Mean<br>Diff<br>(%) | p-value* |
| Haemoglobin (g/L) | 121.1<br>(13.5) | 123.6<br>(12.0) | 2.55<br>(2.1%) | 0.27 | 115.0<br>(16.6) | 115.5<br>(15.3) | 0.46<br>(0.40%) | 0.31 |
| White blood cells<br>($\times 10^3$ /mCL) | 6.0<br>(2.6) | 7.4<br>(5.7) | 1.38<br>(23.1%) | >0.99 | 6.2<br>(2.3) | 6.6<br>(3.2) | 0.38<br>(6.1%) | 0.77 |
| Platelets ($\times 10^3$<br>/mCL) | 236.4<br>(149.4) | 250.4<br>(159.4) | 14.00<br>(5.9%) | 0.17 | 215.2<br>(76.5) | 235.7<br>(116.2) | 20.54<br>(9.5%) | 0.46 |
| eGFR<br>(mL/min/1.73m<sup>2</sup>) | 97.0<br>(36.9) | 94.3<br>(32.0) | -2.73<br>(-2.8%) | 0.71 | 103.6<br>(25.5) | 104.3<br>(24.7) | 0.73<br>(0.7%) | 0.82 |
| Alkaline<br>phosphatase (IU/L)<br>Missing data =2 | 112.4<br>(42.7) | 133.7<br>(65.4) | 21.36<br>(19.0%) | 0.20 | 160.3<br>(98.0) | 164.6<br>(102.6) | 4.38<br>(2.7%) | 0.76 |
| CEA level (ng/mL)<br>Missing data =5 | 15.6<br>(32.4) | 13.7<br>(20.4) | -1.93<br>(-12.4%) | 0.81 | 83.4<br>(184.1) | 49.3<br>(99.2) | -34.10<br>(-40.9%) | 0.71 |
| Lactate<br>dehydrogenase<br>(mmol/L)<br>Missing data =10 | 230.9<br>(121.0) | 180.6<br>(40.7) | -50.29<br>(-21.8%) | 0.44 | 299.8<br>(169.4) | 252.7<br>(93.4) | -47.05<br>(-15.7%) | 0.26 |

\* Between means (Wilcoxon matched-pairs signed-rank test)

**Figure-1:**
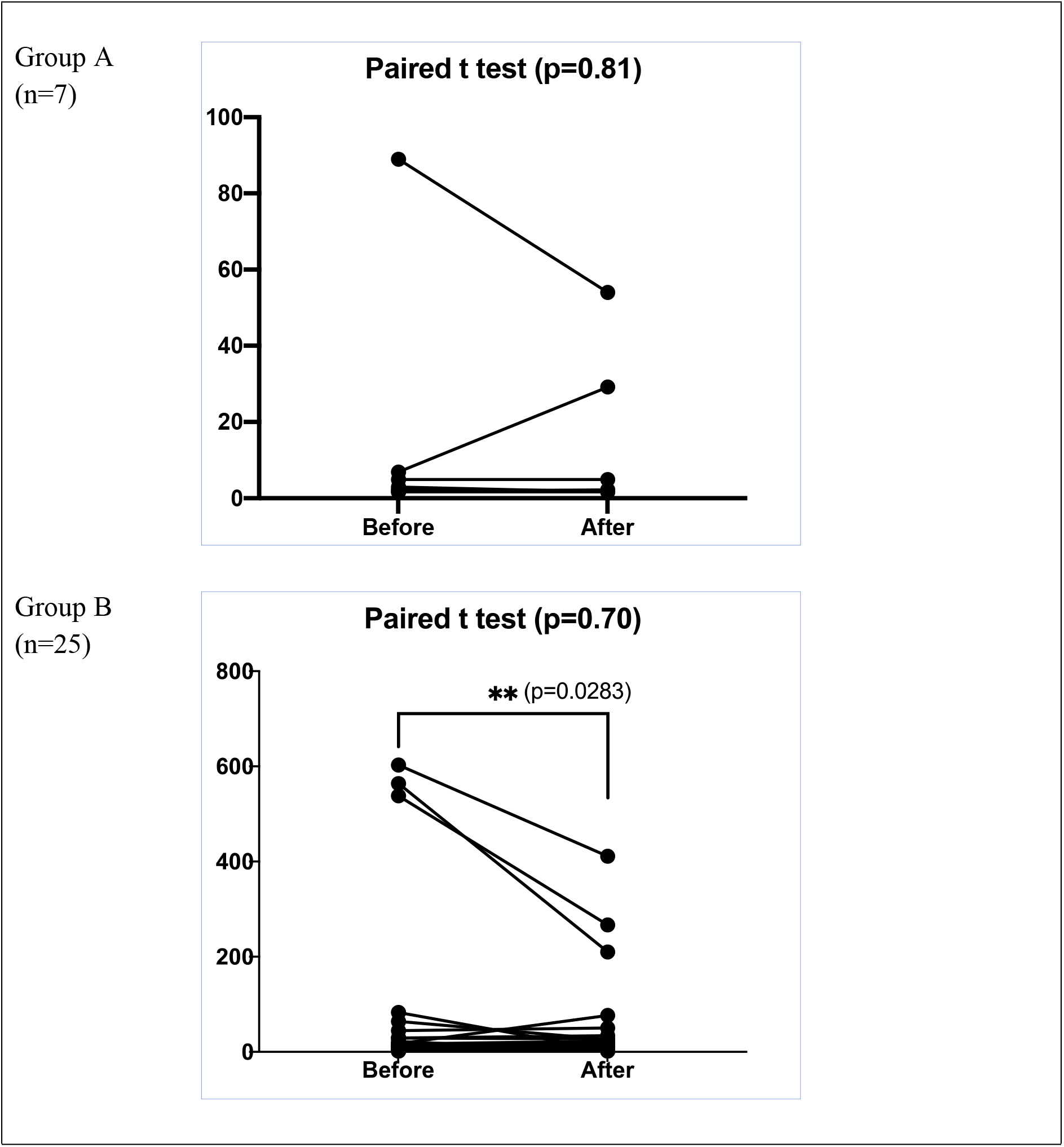
CEA levels (ng/mL) before versus after the end of the month of Ramadan in the low-fasting (Group A) and the high-fasting (Group B). The p-value (0.0283) is calculated for only the three samples that showed a significant reduction in CEA levels.

## Discussion

Despite the notable advances in cancer treatment, cancer remains a considerable burden on the healthcare system. According to the Global Cancer Statistics 2020 (GLOBOCAN), about 19.3 million new cancer cases were estimated Worldwide in 2020, with 28.4 million new cases by 2040 [8]. CRC is among those commonly diagnosed malignancies among adults in men and women. CRC is the third most common cancer in the US after lung and breast cancer, with incidence rates of 30% higher in men than in women [11]. Meanwhile, the overall death rate due to CRC has continued to decline over the last few decades reflecting improvement in the monitoring and treatment of CRC [8].

Family history and screening of intestinal adenomatous polyps before they become cancerous are pivotal for the early detection of CRC. It is usually challenging to recognize such carcinoma because it does not produce definitive classical symptoms. According to the American Society of Clinical Oncology and the European Society for Medical Oncology guidelines, regular screening of asymptomatic adults with an average risk is recommended at 50 years old [12, 13]. This age has decreased recently to 45 years due to the steady increase of CRC amongst the young. However, there are few sensitive and specific tumor biomarkers for detection and/or assessing the efficiency of anticancer therapy. Currently, tumor biomarkers are rarely used to screen cancer either due to the absence of specific biomarkers or their inconsistent expression in cancer cells. Serum carcinoembryonic antigen (CEA) is the most widely known diagnostic and prognostic tumor biomarker in recurrent CRC [14]. Lactate dehydrogenase (LDH) has also been utilized in the oncology practice as a biomarker of the tumor prognosis [15]. Both biomarkers are not specific for CRC only, as elevated levels of CEA and LDH can also be concurrent with other cancers and in several inflammatory and metabolic disorders [16, 17]. Nevertheless, CEA remains a non-invasive tumor biomarker to monitor CRC progression and the efficacy of anticancer treatment [18].

The current study investigated changes in the blood parameters and levels of the two tumor biomarkers [CEA and LDH] due to intermittent fasting during the month of Ramadan in CRC patients receiving chemotherapy. The results did not reveal any significant difference in the measured variables between the initial values and by the end of the 30 days of the month of Ramadan. Although statistically insignificant, the levels of CEA and LDH were reduced in the majority of CRC patients. However, the mean level of CEA in the fasting group was substantially reduced by more than 40% attributed to the highly significant decline of CEA levels in three patients only (p=0.0283) who had remarkably extreme levels of CEA. Nonetheless, the high CEA levels were associated with poor survival in CRC patients [19]. Several studies have appraised the role of CEA tumor biomarker as a diagnostic and prognostic tool, especially in the advanced CRC stages [14, 16, 20]. Others reported suboptimal accuracy of this tumor biomarker for CRC [21, 22].

On the other hand, quite a few studies have investigated the role of LDH, a key enzyme in glycolysis, as a prognostic tumor biomarker in clinical practice [17]. High LDH levels are usually associated with significant tumor burden due to the invasiveness and chemotherapy resistance in several malignancies, including CRC [23, 24]. A recent study has employed the LDH-to-albumin ratio as a prognostic biomarker in CRC patients, which was associated with poor prognosis in CRC patients [25]. Results of the current study have revealed an insignificant decrease in the levels of LDH among CRC patients in both fasting and non-fasting groups. However, some studies reported a significant correlation between the LDH levels and chemotherapy response as well as with the overall survival in patients with high LDH levels [26, 27]. Other studies reported significant benefits of bevacizumab as a targeted therapy, when combined with the standard of care treatment of metastatic and advanced CRC in patients with high LDH levels [28].

Therefore, all these results support the potential role of CEA and LDH levels as predictive tumor biomarkers in CRC patients. Moreover, the combined assessment of these tumor biomarkers, with possibly other parameters [29], rather than relying on a single element, can facilitate the early detection of CRC recurrence, besides their utilization as a prognostic tool and eventually during the follow-up of anticancer therapy.

This study also investigated the potential benefits of intermittent fasting in CRC patients receiving chemotherapy during the month of Ramadan. The majority of patients in the current study have reported improvement in intolerability of the chemotherapy side effects. Less than 20% of patients reported worsening nausea while fasting, but it did not affect their usual activities of daily living. This is consistent with a number of studies that have reported no significant changes during short-term fasting [30, 31] or even a substantial reduction in the chemotherapy-related adverse effects, particularly in hematological toxicity [32], GI upsets [33] and fatigue [34]. Due to the hot and dry climate, dehydration and the renal and liver status are among the critical concerns of prolonged fasting periods in the current study settings. However, both eGFR and alkaline phosphate showed no significant changes, minimizing such concerns.

Moreover, several reports have also suggested a potential improvement in the cancer therapeutic outcomes if cancer chemotherapy was combined with short-term fasting due to restriction of nutrient intake without causing weight loss [35]. Furthermore, fasting could even augment cancer immunotherapy strategies by inducing cancer-specific T cell activation, which plausibly enhances the cancer cell killing effect [36].

All these previous results have confirmed the safety and the potential benefit of intermittent fasting or short-term fasting during chemotherapy due to alleviation of adverse effects and possibly attributed to the profound metabolic changes in the human body that occurs during nutrient restriction such as reduction of the circulating levels of growth factors, and inflammatory cytokines that are associated with various malignancies [37, 38].

Lastly, the small sample size is an inevitable limitation of the current study limitations that may have impacted the significance and generalizability of the study results. Furthermore, it is important to report that CEA and LDH levels are not routinely measured during the adjuvant chemotherapy setting in CRC patients. Recruited patients in this study had different CRC stages, some being in the early stage (II and III) and others being in the late stage IV. Hence, the effects of fasting on CEA and LDH levels may perhaps be overestimated.

## Conclusion

The results of this study have been consistent with several reports that have demonstrated the safety of intermittent fasting with enhanced tolerability and possibly improved therapeutic outcomes of cancer chemotherapy. Nonetheless, limited data explored the impact of fasting in cancer patients before verifying the potential health benefits. The levels of CEA and LDH tumor biomarkers, which are frequently associated with CRC, were not valuable in assessing the benefits of intermittent fasting during chemotherapy. Therefore, further studies with a larger population and more extended follow-up periods are still necessary to evaluate the role of CEA and LDH levels as screening, diagnostic, and prognostic tumor biomarkers or to monitor patients’ responses to anticancer therapy.

## Data Availability

Summary of data is provided in the manuscript. Detail data of patients cannot be shared due to confidentiality and privacy of patients' data. Cases that are enrolled in the study are going to be identified by anonymous serial numbers. The study team will follow the regulations of the KAIMRC and the GCP-ICH standards.

## Acknowledgments

The authors would like to thank the whole team in the Oncology Department at KAMC and KASCH for their indispensable assistance during data collection.

